# Inter-articulatory Timing of Vocalic Tongue Body and Jaw Movements across Talkers with Dysarthria due to Various Neurological Conditions

**DOI:** 10.64898/2026.11.13.26368039

**Authors:** Tabea Thies, Mary S. Dietrich, Antje Mefferd

**Affiliations:** Department of Neurology, Faculty of Medicine and University Hospital Cologne, University of Cologne, Cologne, Germany; IfL Phonetics, Faculty of Arts and Humanities, University of Cologne, Cologne, Germany; Department of Hearing and Speech Sciences, Vanderbilt University Medical Center, Nashville, TN, USA; School of Nursing, Vanderbilt University, Nashville, TN, USA; Department of Biostatistics, Vanderbilt University Medical Center, Nashville, TN, USA

**Keywords:** phase angle, articulatory timing, dysarthria, speech kinematics, phasing

## Abstract

**Purpose:** This study compared inter-articulatory timing patterns of the tongue and jaw across talkers with Parkinson’s disease (PD), amyotrophic lateral sclerosis (ALS), Huntington’s disease (HD), multiple sclerosis (MS), and neurotypical controls (CON) to identify disease-related differences. The study also explored if a talker’s tongue body cycle duration, which serve as a proxy for a talker’s habitual speech rate, or a talker’s reliance on their jaw, as indicated by their jaw movement amplitude, could predict their inter-articulatory timing patterns.

**Method:** Using three-dimensional electromagnetic articulography, the movements of the posterior tongue and jaw were examined in 24 talkers with PD, 19 talkers with ALS, 18 talkers with HD, 18 talkers with MS, and 34 CON during a sentence repetition task (“Buy Kaia a kite”). The tongue movement during the segment [kai] served as the reference signal (tongue body cycle) for which the phase angle of the jaw reaching the maximum opening position during [kai] was calculated. Tongue body cycle durations were measured as the time between the tongue body movement onset to the tongue body movement offset. The jaw opening amplitude was measured as the relative positional change from the jaw’s maximum closing position associated with [k] to the jaw’s maximal opening position at the onset of the diphthong [ai].

**Results:** A significant group effect was found for phase angles. Pairwise comparisons showed that talkers with ALS had significantly larger phase angles than talkers with PD, MS, HD, and CON, indicating a timing pattern where the tongue reached the target [a] earlier relative to the jaw. No significant differences were found for any other pairwise comparisons. Furthermore, no significant associations were found between tongue body cycle durations and phase angles. A significant association was found between jaw amplitudes and phase angles when considering all talkers [*r_S_*(110) = .253, *p* = .008] and when only considering talkers of the control group [*r*(34) = .361, *p* = .036].

**Conclusions:** Findings suggest that during the production of [kai], tongue-jaw timing patterns of talkers with ALS differ from those of talkers with PD, MS, HD, and CON with the tongue reaching the maximum opening position before the jaw while the opposite pattern was observed for talkers of the other groups. However, the variability in tongue body cycle duration did not significantly contribute to the variability in phase angles and the variability in jaw amplitudes only accounted for 6% of the variability in phase angles across all talkers. Future studies are needed to determine if the observed tongue-jaw timing pattern of talkers with ALS generalizes to other phonetic contexts and determine the underlying factors that contribute to the unique tongue-jaw timing patterns of talkers with ALS.

## Introduction

Speech production is a complex goal-directed motor behavior that requires a smooth interaction of multiple speech subsystems (i.e., respiration, phonation, resonance, and articulation). At the articulatory level, motor execution is carefully timed across the tongue, upper and lower lips, and the jaw, to achieve the desired acoustic output [1–3]. These inter-articulatory timing patterns are thought to play a critical role in the production of intelligible speech [4,5]. They can also provide insights in the spatiotemporal programming and coordination strategies of the motor control system. In this study, inter-articulatory timing is understood as a specific, quantifiable aspect of speech motor coordination that describes how distinct speech structures are spatiotemporally aligned. While closely linked to coarticulation, the context-specific physiological overlapping of adjacent speech gestures executed by one articulator, inter-articulatory timing focuses precisely on the coordination between two or more articulators. Inter-articulatory timing patterns can be quantified by relative phasing, expressed as the positional and spatiotemporal state of one articulator relative to another articulator at a specific landmark in terms of a phase angle [6–8] or temporal latencies, expressed as the absolute time difference between articulatory events (velocity peaks, positional targets) of two articulators [8,9]. This work focuses on the former, quantifying the relative phasing of the jaw relative to the tongue via phase angles.

### Phase angles

In previous studies on neurotypical talkers, phase angles have been used to examine spatiotemporal relations between two articulators. In such analyses, the movement of one articulator (typically the jaw) served as the reference signal, to which lip or tongue tip movements were related during consonant-vowel-consonant sequences such as [bab] or [tat] [10,11]. The rationale for a target utterance with two stop consonants was that it provided robust discrete kinematic landmarks to define the movement cycles. The calculation of phase angles is based on the concept that articulators are oscillators producing sinusoidal-like movement cycles along a phase plane [6]. Consequently, the relative timing between two articulators can be quantified as phase angles. In this framework, a phase angle of 180 degrees signifies a synchronized timing pattern. Phase angles less than 180 degrees indicate that the reference articulator lags behind the second articulator, while angles greater than 180 degrees suggest that the reference articulator leads the second articulator. This approach provides a nuanced representation of the spatiotemporal relationship between articulators during speech production (Fig. 1).

**Fig 1:**
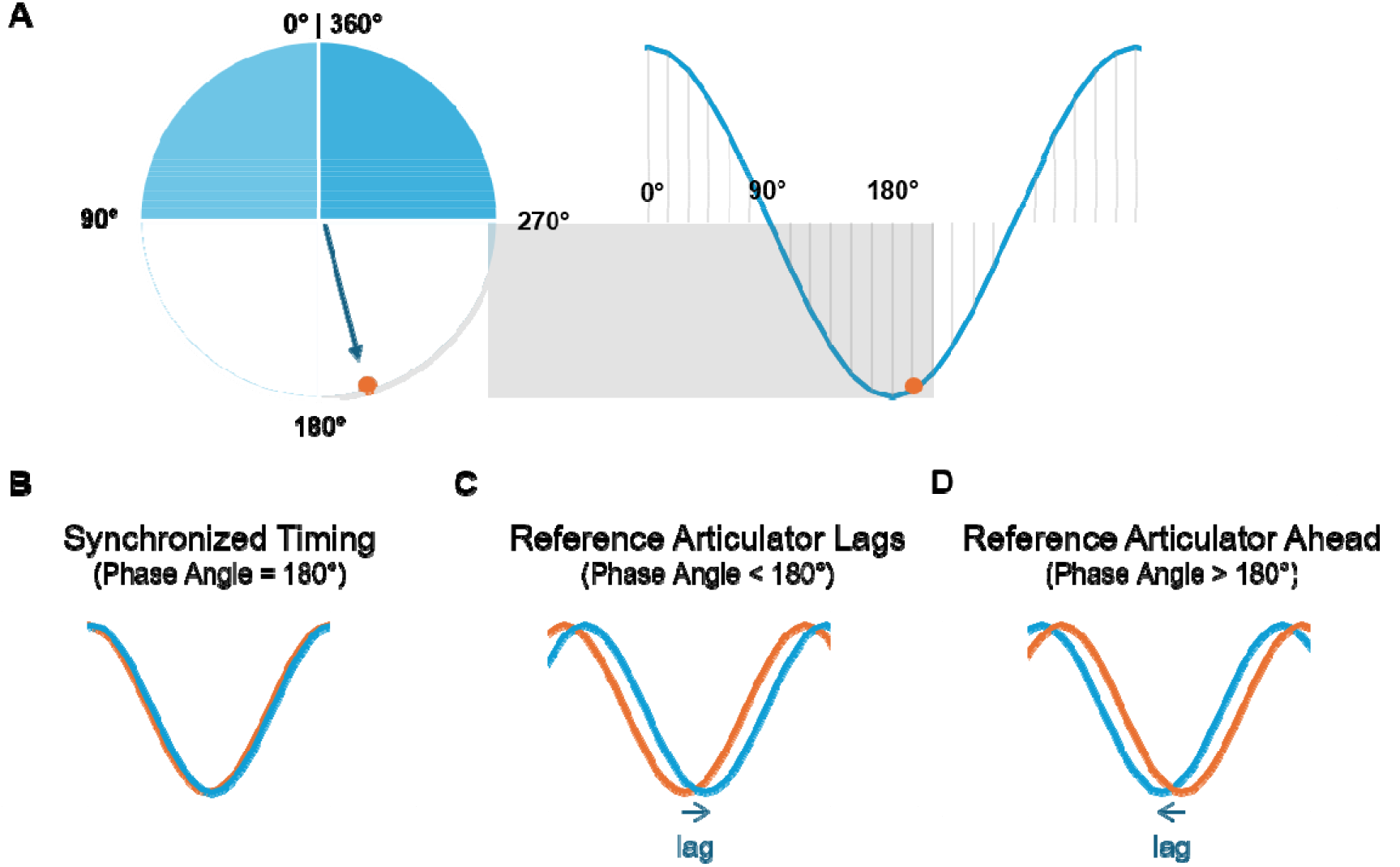
Conceptualizing Inter-Articulatory Timing via Phase Angles. A: The reference articulator as an oscillator along a phase plane. B to D: Relative timing between two articulators. Reference articulator in blue, related articulator in orange.

In their seminal work, Tuller and Kelso [12] examined inter-articulatory timing patterns of the upper and lower lip movements relative to the jaw cycle and reported relatively stable phase angles that were not affected by changes in jaw cycle duration (which were typically used as a proxy for quantifying a change in speaking rate or lexical stress). By contrast, the majority of studies conducted later on suggested that the timing patterns do change when talkers manipulate their lexical stress patterns or speech rate [7–9,13,14]. Specifically, speech demands that resulted in a shortening of the jaw cycle duration (e.g., fast speech, unstressed segments) also decreased the phase angle. Along those lines, speech demands that facilitated longer jaw cycle durations (e.g., slow speech, stressed segments) were associated with an increase in the phase angle, given that movement cycle duration serves as an established proxy measure for speaking rate. However, a great deal of variability was noted across talkers and across different articulator pairs [12,15]. Specifically, while upper lip timing maintained a relatively stable phase angle range relative to the jaw, tongue tip phase angles exhibited much greater fluctuations [7,13]. Nevertheless, tongue tip phasing remained sensitive to rate, consistently varying with jaw cycle duration despite a greater underlying variability.

### Temporal latencies

Temporal latencies refer to the temporal lag between kinematic trajectory landmarks purely in the time domain, independent of the articulators’ positions and velocities. Previous studies investigated tongue tip - jaw and lower lip - jaw timing patterns during the production of disyllabic utterances in neurotypical talkers [9, 10, 11, 16, 17]. In these studies, temporal latencies were measured as the time elapsed from the onset of the jaw vowel cycle until the initiation of the upward movement (raising) of the tongue tip or lower lip toward the intervocalic consonant [9,11]. An important finding of these studies was that a shortening of the jaw opening-closing cycle due to speaking rate or lexical stress manipulations systematically reduced the movement onset latency of the second articulator as well. These precise temporal regularities held true across different sets of articulators (tongue-jaw and lip-jaw synergies), which suggests that synergistic timing patterns of vocal tract constriction formations are precisely controlled regardless of where in the vocal tract the constriction occured [10,16,17].

### Inter-Articulatory Timing Patterns in Talkers with Dysarthria

Only a limited number of kinematic studies are currently available that have examined inter-articulatory timing patterns and the majority of them investigated neurotypical talkers. However, it is commonly assumed that inter-articulatory timing patterns are disrupted in talkers with dysarthria, a neurogenic motor speech disorder that arises when one or more neural components of the speech motor system are affected by a neurological condition. Rather than presenting as a uniform deficit, dysarthria encompasses distinct subtypes depending on the neural components that are affected. In amyotrophic lateral sclerosis (ALS), for example, upper and lower motoneurons of bulbar and spinal muscles degenerate, which results in a mixed flaccid-spastic dysarthria. Hallmark symptoms are an abnormally slow speaking rate, imprecise consonants and distorted vowels, and a strained-strangled voice quality [18,19]. Multiple sclerosis, by contrast, commonly affects the corticobulbar tract and cerebellar motor control circuits, which can result in a mixed dysarthria with spastic and ataxic components. Common perceptual speech characteristics of talkers with MS include a slow speaking rate, irregular articulatory breakdowns, consonant imprecision, and a breathy or harsh voice quality [18,20]. In other diseases, such as Parkinson’s disease (PD) and Huntington’s disease (HD) basal ganglia control circuits are commonly affected. PD typically results in hypokinetic dysarthria, which is associated with an accelerated speech rate, short rushes of speech, reduced vocal loudness or loudness decay, a breathy or harsh voice quality, and consonant imprecision [20]. Finally, speech disturbances in Huntington’s disease (HD) typically manifest as hyperkinetic dysarthria and are characterized by involuntary movement intrusions, a variable speech rate, consonant imprecision, and irregular articulatory breakdowns.

Only few kinematic research studies are currently available on inter-articulatory timing patterns in neurogenic populations, such as individuals with PD and ALS. These studies have provided crucial insights into how two neuropathologies can differentially alter inter-articulatory timing patterns [21,22]. Specifically, Weismer et al. (2003) investigated labial and lingual timing patterns by means of temporal latencies. They reported that overall timing patterns of the tongue and lips remained largely unaffected in PD; however, individuals with ALS exhibited a significantly wider range and greater disruption of lip and tongue timing patterns [22]. Extending these observations to a spatiotemporal metric, Rong and Heidrick (2021) found that relative phase relationships between the tongue and jaw served as robust markers of bulbar involvement, with relative phasing achieving high diagnostic accuracy in differentiating individuals with ALS from healthy controls [21].

Beyond neurodegenerative conditions, there is only one kinematic study currently available that examined inter-articulatory timing patterns in talkers with dysarthria resulting from traumatic brain injury. That study found no significant group-level differences in inter-articulatory timing patterns between nine talkers with traumatic brain injuries and nine typical talkers [23]. Nevertheless, an inspection of individual performance patterns revealed disrupted inter-articulatory timing patterns in those talkers with traumatic brain injury who displayed the most severe dysarthria symptoms. Given the small sample size, this study offers very limited insights into how inter-articulatory timing patterns vary more broadly across of talkers with dysarthria.

Given the limited kinematic literature on speech motor control in talkers with dysarthria, the current study sought to investigate inter-articulatory timing patterns of the tongue and the jaw in talkers with dysarthria with different neurological conditions [i.e., PD, MS, ALS, HD]. The comparisons across these four disease types are particularly interesting and important because these diseases are associated with different neuropathologies (i.e., differential disruptions to the basal ganglia control circuits in PD versus HD; disruptions to the direct motor pathway in MS and ALS, but additional cerebellar control circuit disruptions in MS). Thus, these four different neurological conditions may give rise to disease-specific deviancies in inter-articulatory timing patterns. It is especially important to investigate the inter-articulatory timing patterns of the tongue body and jaw in these talkers because tongue and jaw motor performance deficits have been reported in talkers with PD, HD, MS, and ALS [24–30]. For example, talkers with ALS are thought to produce excessive jaw movements to compensate for impaired tongue muscles during speech production. An overreliance on the jaw may result in more synchronized tongue and jaw movements in these talkers as indicated by phase angles close to 180 degrees. By contrast, talkers with MS, PD, and HD may exhibit less synchronized tongue and jaw movements considering that lesions in cerebellar and basal ganglia motor control circuits have been associated with the disruptions of temporal patterns during speech production [26,30]. Thus, these talkers may have phase angles that are below or above 180 degrees.

However, it is also possible that inter-articulatory timing patterns of talkers with MS and ALS differ from those of talkers with PD due to differences in their speaking rate. Based on the known speech motor profiles of different dysarthria subtypes, distinct timing disruptions may exist. That is, talkers with PD may exhibit inter-articulatory timing patterns that mimic the timing patterns of healthy controls when asked to speak at an accelerated rate (fast speech), while talkers with MS and ALS may exhibit timing patterns that mimic those of controls when asked to speak at a reduced rate (slow speech). Finally, the speech rate of talkers with HD is commonly described as variable [24], which may disrupt inter-articulatory timing patterns of these talkers in unpredictable ways.

In sum, the aims of the current study were:

- Aim 1: To characterize speaking rate of talkers with dysarthria, we determined between-group differences in tongue body cycle durations across clinical groups and controls
- Aim 2: To characterize the reliance on the jaw in talkers with dysarthria, we determined between-group differences in jaw amplitudes across clinical groups and controls
- Aim 3: To characterize inter-articulatory timing patterns of the tongue and jaw in talkers with dysarthria, we determined between-group differences in phase angles across clinical groups and controls
- Aim 4: To determine if differences in speaking rate account for differences in inter-articulatory timing patterns in talkers with dysarthria, we evaluated the association between tongue body cycle durations and phase angles.
- Aim 5: To determine if differences in the reliance on the jaw account for differences in inter-articulatory timing profiles in talkers with dysarthria we evaluated the association between jaw amplitude and phase angle.

## Methods

### Participants

The experiment took place at the Vanderbilt University Speech Kinematics and Acoustics Lab. The study was approved by the Vanderbilt University Medical Center IRB (IRB #150655). All participants provided written consent prior to data collection and were compensated for their time. All participants were recruited as part of a larger ongoing project during the period from 08/08/2015 to 06/24/2025.

The current study included 19 talkers with ALS, 19 talkers with MS, 18 talkers with HD, 24 talkers with PD, and 34 CON. Demographic information for each clinical group and CON can be found in Table 1. All participants were native speakers of American English. Talkers with dysarthria were diagnosed with either PD, HD, ALS, or MS by a board-certified neurologist and reported no comorbid neurological conditions. CONs had no history of a neurological condition per self-report. Participants reported no previous history of a speech, language, or hearing impairment. Participants completed either the Mini-Mental State Examination (MMSE) [31] or the Montreal Cognitive Assessment (MoCA) [32]. The cognitive screening was used to ensure that participants were able to follow instructions and complete the task during the experiment. Participants scored at least 20 points on the MMSE or the MoCA.

**Table 1.**
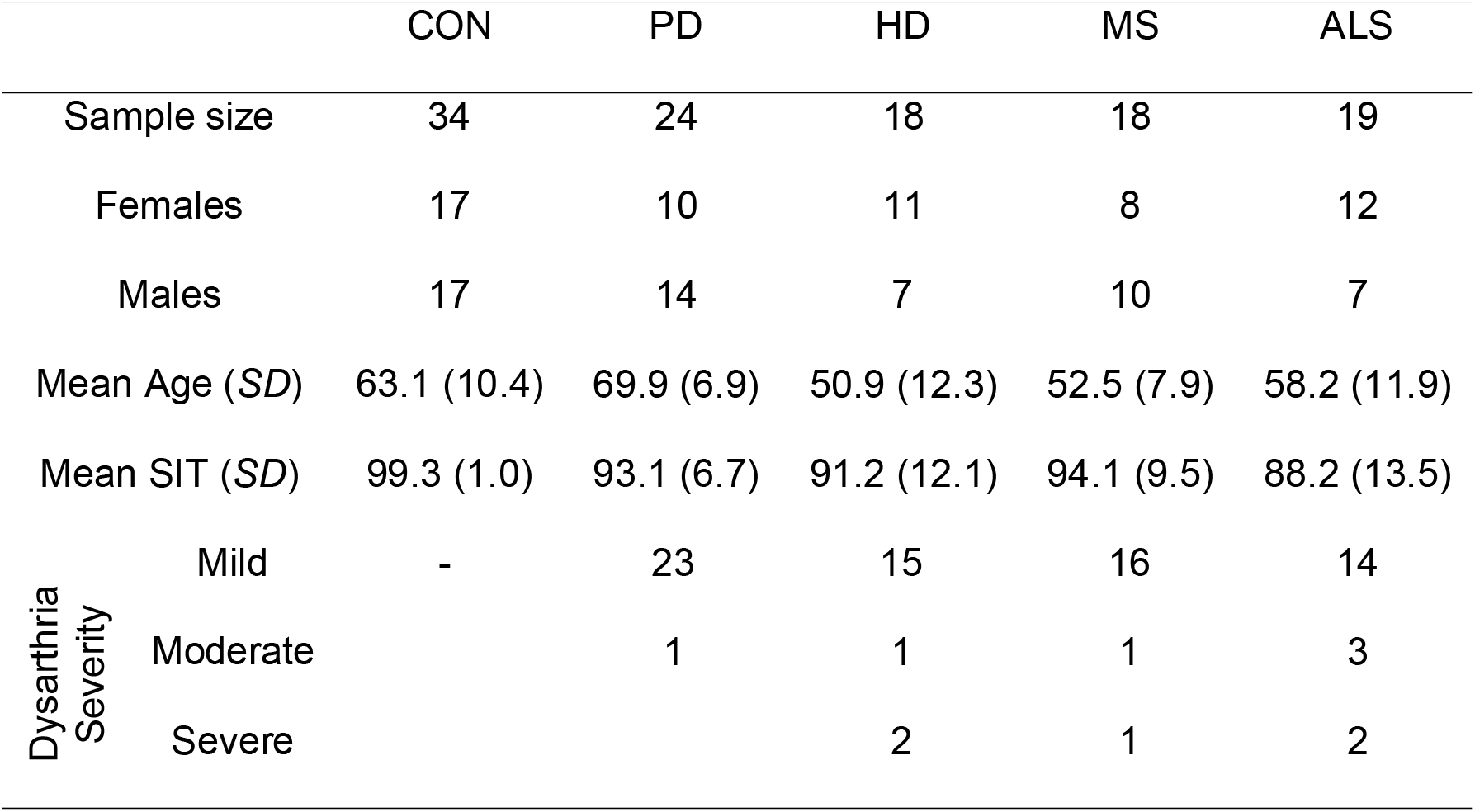
Participant demographics.

All participants completed the Sentence Intelligibility Test (SIT) [33] with a new set of sentences generated for each participant. Each participant’s recording of the SIT was orthographically transcribed by three student research assistants. They were asked to listen no more than twice to each sentence and transcribe it using the SIT computer software. Speech intelligibility was calculated as the percentage of words correctly transcribed out of the total number of words elicited during the test. A mean SIT score was calculated for each participant based on the SIT scores of the three student research assistants. SIT scores for each clinical group and typical talkers can be found in Table 1. SIT scores were used to determine the dysarthria severity of the participants in each clinical group. We used cut-off scores of 85%, 70%, and 50% for mild, moderate, and severe dysarthria, respectively. There was one talker with PD, for whom a SIT score could not be obtained because of a technical error with the audio recording. This talker’s dysarthria severity was based on a paragraph reading task. Dysarthria severity distributions for each clinical group can be found in Table 1.

All talkers with dysarthria presented with a perceptible articulatory subsystem impairment as indicated by perceptible imprecise speech, phoneme prolongations, phoneme repetitions, and/or an abnormal speaking rate during at least one of the following speech tasks: SIT, paragraph reading, or picture description. The presence of an articulatory subsystem impairment was determined based on consensus ratings of two speech language pathologists (SLPs, one being the second author). In five cases, a third SLP was included to reach a consensus.

All participants with PD and six out of 18 participants with HD were on medication that aimed to reduced motor-related disease symptoms at the time of data collection. Three participants with PD had been treated surgically with deep brain stimulation (DBS) and were recorded while the device was turned on.

### Speech Material and Data Collection

Kinematic speech data were recorded with an electromagnetic articulograph (AG 501; Carstens Medizinelektronik GmbH). Small sensors were attached to the articulators to track their movements during speech. That is, one sensor was attached with a small amount of Stomahesive® putty to the gumline of the lower central incisors to track jaw movements. Additional sensors were placed at midline on the vermillion border of the upper and lower lips using dental adhesive (Periacryl 90, GluStitch). Two sensors were placed with dental adhesive on the tongue. One tongue sensor was attached approximately 1 cm posterior to the tongue tip to track its movements. The other tongue sensor was placed approximately 4 cm posterior to the tongue tip to assess tongue body movements. Three additional sensors were placed on plastic goggles (head right, head center, head left) that the participants wore during data collection. These sensors served as reference sensors for head correction [28]. Finally, three sensors were placed on a biteplate, which participants were asked to hold between their front teeth to record a short rest file.

After attaching all sensors, participants were engaged in conversations to provide them the opportunity to familiarize themselves with the presence of the sensors in the oral cavity. Afterwards, the experimental protocol was started. As part of a larger experimental protocol, participants were asked to produce the sentence “Buy Kaia a kite”, which was displayed on a computer monitor in front of them. All participants were asked to repeat the sentence ten times at their typical speaking rate. “Kaia” was produced as /’kai. ə/. If a participant did not produce the word “Kaia” correctly, the examiner modeled the correct pronunciation and ask the participant to repeat the task.

### Data processing

The raw data were converted into positional data using the CalcPos software (Carstens Medizintechnik GmbH). Then all positional data were head-corrected, down-sampled to 250 Hz, and transposed into a head-based coordinate system using the NormPos software (Carstens Medizintechnik GmbH). Data were filtered with a low-pass filter at 15 Hz and displayed and further processed in SMASH, a MATLAB-based software program [34]. The data analysis focused on the first, stressed syllable of the word “Kaia”, consisting of a stop consonant and a diphthong (/’kai./). For the purpose of this study, only the tongue body and jaw movement in the vertical dimension (i.e., y-dimension) were considered.

Within the movement sequence of the tongue body during the syllable [kai], the onset of the jaw raising movement during the diphthong [ai] can be determined as an angle on the tongue body’s phase plane. The segment [kai] was of particular interest in this study because, although the tongue body and jaw are anatomically coupled, the tongue and the jaw can independently reach the maximum vocal tract opening at the onset of the diphthong different times and, consequently, initiate the movements towards the maximum vocal tract closing at the offset of the diphthong at different times. Furthermore, the inter-articulatory timing patterns of the jaw and tongue body were also of interest because the velar place of articulation has been shown to be affected at the early stages of articulatory performance decline in talkers with neurodegenerative diseases [35].

We did not decouple the jaw from the tongue in the current study. An estimation of independent tongue movement based on linear subtraction of the jaw movement would have resulted in an overestimation of the jaw contribution due to the jaw sensor’s location being more anterior from the center of the jaw rotation than the posterior tongue sensor’s location [36]. Consequently, the phase angles reported here describe the relationship of the jaw relative to the tongue body+jaw composite movement rather than to an independently derived tongue body movement. While individual differences in the intrinsic contribution of the jaw to tongue movement could not be fully captured without decoupling, pilot trajectory inspections confirmed consistent asynchrony between jaw and tongue positional minima. Thus, the phase angles in this study represent only the relative phasing of the jaw in relation to the tongue body+jaw movement during the syllable [kai].

Contrary to most previous studies in which the jaw cycle was used as a reference, in the current study the reference for the phase angle calculation was the tongue body cycle. For the selected target word, the jaw could not be used as a reference because its movement did not offer a reliable landmark for the cycle offset. Instead, the vertical movement of the tongue body during the sequence [kai] offered reliable kinematic landmarks. The local maxima in the tongue movement were associated with the constriction of [k] and the maximum closing at the offset of the diphthong [ai] and the local minimum was associated with the maximum vocal tract opening at the onset of the diphthong [ai]. However, for two talkers with ALS and one talker with HD, the tongue body movement of all ten sentence repetitions did not contain the second local maximum. Another talker with HD could only produce the second local maximum in two of ten sentence repetitions. When the second local maximum was missing, the diphthong was either severely distorted or replaced by the vowel [a] and a glottal stop. In these cases, phase angles could not be calculated.

Using SMASH [34], the vertical tongue movement and the vertical jaw movement were trimmed using the local maxima the tongue body associated with [k] and the offset of the diphthong [ai] as the onset and offset of the tongue body cycle. These local maxima were algorithmically determined in SMASH based on the zero crossing of the velocity signal of the vertical tongue movement (see also Fig 2). In instances where multiple local maxima were observed, the last local maximum before the opening movement was selected for the onset and the first local maximum after the closing movement was selected for the offset. The trimmed trajectories of the tongue and jaw were saved as a new file. A custom-written Matlab script was then used to calculate the velocity signal of the trimmed tongue body movements. To ensure precise temporal alignment when extracting the tongue velocity at the time of maximum jaw opening, the MATLAB script employed central differencing via the gradient function to avoid time-shift artifacts associated with the standard diff function that is used in SMASH.

**Fig 2.**
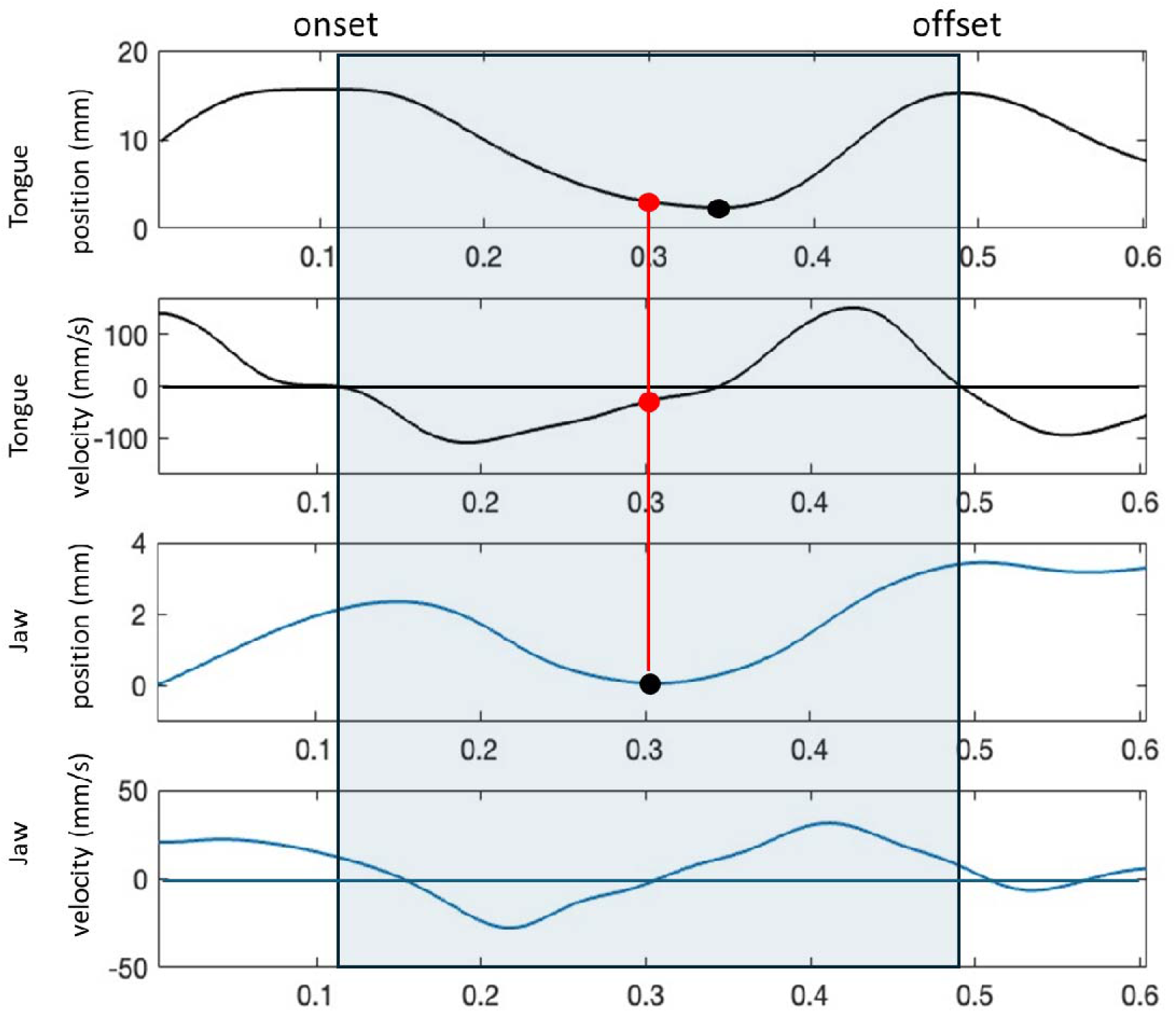
Vertical tongue and jaw movement trajectories and their corresponding velocity signals during the sentence “Buy Kaia a kite” based on a female neurotypical talker. Vertical black lines indicate the onset and offset of the target segment [kai] (shaded) based on the zero crossing of the velocity signal of the tongue body. The black circles mark the positional minimum during the maximum opening of the tongue and jaw at the diphthong onset. After spatial normalization of the tongue position and velocity signal, the normalized tongue body position and velocity value at the time of the jaw reaching the positional minimum (marked with red circles) was extracted to calculate the phase angle.

To compute phase angles, position and velocity signals were mapped onto a phase plane. Because position (mm) and velocity (mm/s) rely on different physical units, both variables were normalized to the numerical interval [−1, +1] within each movement cycle prior to applying the inverse tangent function, following the classic procedure described by Kelso et al [6]. Thus, the spatial normalization of the tongue body positions was calculated for the complete tongue body cycle while the velocity was normalized on the half-cycle in which the jaw reached the positional minimum at the maximum jaw opening [6]. As a result, the normalized tongue body cycle ranged in values from −1 to 1 with the positional minimum of the tongue at the onset of the diphthong [ai] nearing the minimum of −1 and the onset and offset of the tongue body cycle nearing the maximum of 1 (see Fig 3A). The normalized velocity ranged from 0 to −1 when the jaw reached the maximum opening position before the tongue and from 0 to 1 when the jaw reached the maximum opening position after the tongue (see Fig 3A). The same Matlab script that calculated the tongue velocity signal was used to spatially normalize the trimmed tongue body movement and tongue velocity signal as described in [6], extract the point of maximum jaw opening at the onset of the diphthong [ai], and calculate the resulting phase angle of the jaw relative to the tongue body (Equation 1).

**Fig 3.**
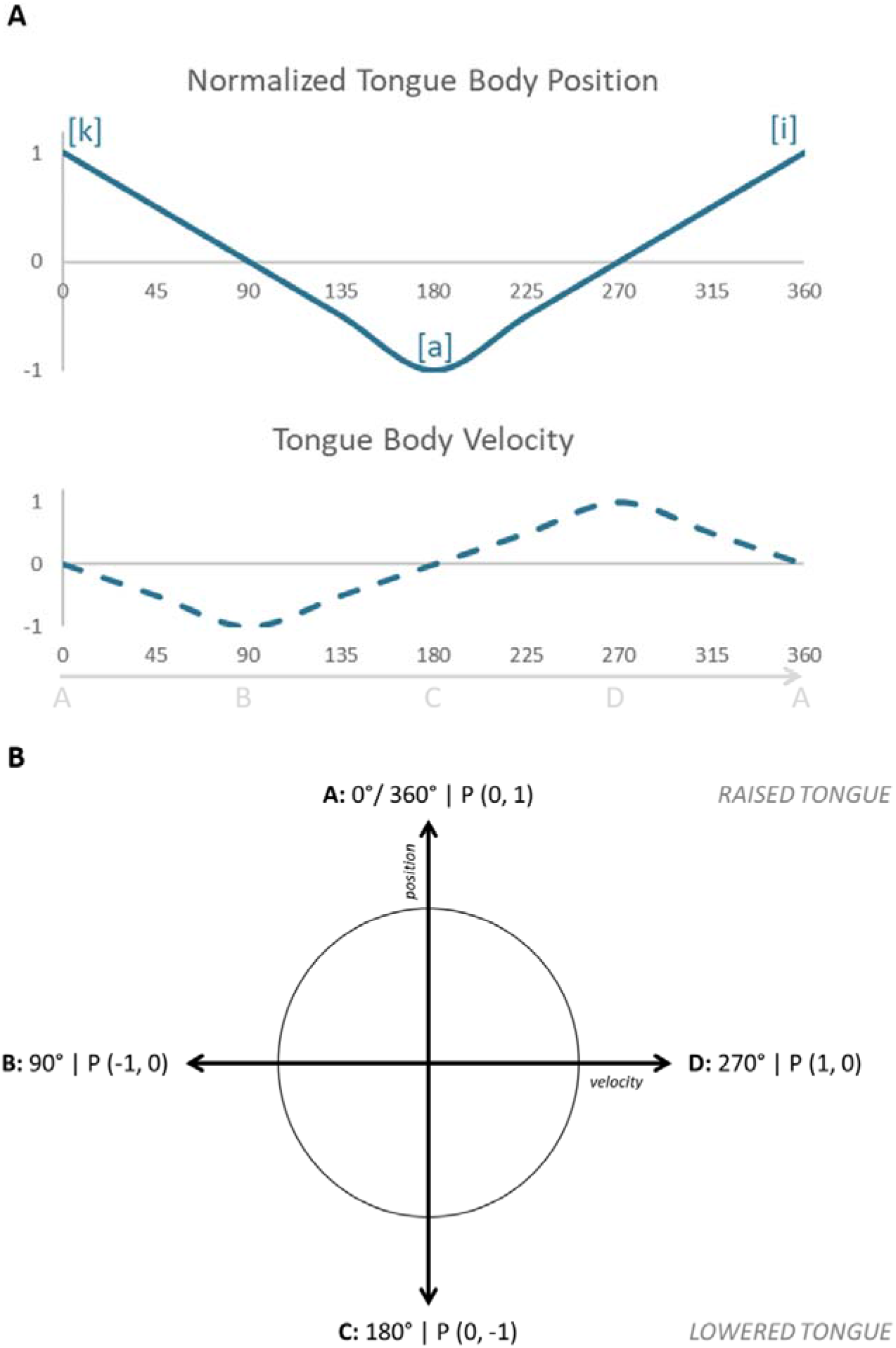
Presentation of one tongue body cycle and its phase plane mapping. Panel A shows the temporal alignment of the normalized tongue body position and the normalized velocity during the production of the target segment [kai]. Panel B illustrates these dynamics as a unit circle (phase plane), where points A–D correspond to specific kinematic events.

A unit cycle is shown in Fig 3B which is centered at P (0,0) and has a radius of 1 defined by normalized position and velocity values. As can be seen in Fig 3B, the tongue position is plotted on the vertical axis while the tongue movement velocity is plotted on the horizontal axis [6]. The points P (velocity, position) describe the movement of the tongue body along the phase plane within the unit cycle. The direction of movement in the current study is counterclockwise. Thus, the tongue body lowering from [k] to the maximum opening position at the onset of the diphthong/ai/ is reflected in the movement from A to C via B (Fig 3B). The velocity is zero at A and C, as the tongue body reaches its highest point P (0,1) and its lowest point P (0,-1), respectively. At B, the tongue body reaches its local velocity maximum as the tongue body reaches its midpoint of the opening phase towards maximum tongue lowering. The tongue body raising from the onset to the offset of the diphthong [ai] is described by the movement from point C to point A via point D. At D, the midpoint of the tongue raising movement, the velocity reaches its local maximum again (Fig 3B).

### Measurements

Using the custom-written Matlab script, the tongue body cycle duration was calculated as an indicator of speech rate as in previous studies, e.g., [11]. Tongue body cycle duration was measured based on the tongue kinematic signal as the time interval between onset and offset of tongue body movement during the target sequence [kai]. Jaw amplitudes were measured as the relative positional change of the jaw from the positional maximum associated with [k] to the positional minimum associated with the onset of the diphthong [ai] during the tongue body cycle. To quantify the inter-articulatory timing pattern of the tongue body and the jaw, the phase angle (φ) was calculated based on the timing of the maximum opening of the jaw relative to the tongue cycle. The local minimum of the jaw was identified algorithmically within the Matlab script. Using the spatially normalized tongue body signal and its first derivative (velocity), the tongue body velocity and the tongue body position at the time of the local minimum of the jaw were extracted. In a few instances the jaw produced multiple local minima. In these cases, the lowest of the local minima was selected to calculate the phase angle. The phase angle of the tongue body was then calculated using Equation 1.

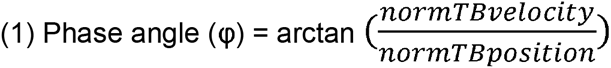

Because the tongue body cycle is used as the reference in the current study, phase angles smaller than 180° indicate that the jaw reached the maximum opening position earlier and initiated the upward movement towards vocal tract constriction sooner than the tongue body. Phase angles larger than 180° indicate that the jaw reached the maximum opening position after the tongue body and initiated upward movement towards vocal tract constriction later. Values of 180° indicate that the jaw and tongue reached their maximum opening positions at the same time and initiated the upward movement towards vocal tract constriction simultaneously.

### Statistical analysis

For each participant, ten repetitions were analyzed unless there were not ten repetitions available due to accidental speech errors (e.g., mispronouncing the target word), miscounting of trials, or missing positional landmarks. In addition to the already mentioned missing second local maximum of the tongue body, there were four instances where the jaw did not produce a local minimum during the tongue body cycle. In these cases, the jaw was continuously raised during the tongue body cycle. These trials were also excluded. Missing or excluded productions were evenly distributed across the cohort rather than clustered in a small number of participants. In total, 1,066 out of 1,100 productions (96.9%) were submitted to the statistical analysis. All statistical analyses were conducted using SPSS Version 32.0.

To select the appropriate statistical tests to assess group effects on tongue body cycle durations, jaw opening amplitudes, and phase angles, data distributions were evaluated for normality, skewness and influential outliers using visual inspections (i.e., histograms and box plots). We found that the distributions for the jaw opening amplitudes were positively skewed. Therefore, we used a generalized linear mixed model (GLMM) with gamma distributions and log link to test for between-group differences in jaw opening amplitudes. In the GLLM we used the residual method to determine the degrees of freedom and a robust estimation to handle violations of the model assumptions.

Phase angle distributions and tongue body cycle durations were not skewed but included some extreme values. Extreme values were defined as values that were more than three times of the interquartile range away from the edge of the first or third quartile (e.g., [37,38]). We removed these extreme values. Specifically, for tongue body cycle durations, we removed one value of one talker with HD and one value for one talker with PD. For phase angles, we removed a total of three values in the control group, six values in the HD group, and one value in the MS group. We then submitted the tongue body cycle durations and phase angles to separate linear mixed models (LMMs). In each model (GLMM, LMM), group was submitted as a fixed factor and repeated trials were submitted as a random factor. If the group effect was found to be significant at *p* < .05, post-hoc pairwise comparisons were completed and the critical alpha level was adjusted for multiple comparisons using Bonferroni corrections.

Because sex was confounded with dysarthria severity in the four clinical groups, sex could not be tested as a covariate in the full models. However, when assessing a potential sex effects in typical talkers, we did not find any significant sex effects on tongue body cycle durations [*F*(1, 334) = 1.806, *p* = .180], jaw opening amplitudes [*F*(1,334) = 1.512, *p* = .220], and phase angles [*F*(1,31.976) = 0.567, *<u>p</u>* =.457].

Similarly, because age was confounded by disease type and dysarthria severity in the current study, age could also not be tested as a covariate in the full models. However, we assessed potential age effects on tongue body cycle durations, jaw opening amplitudes, and phase angles by submitting each control talker’s age and their mean tongue body cycle duration, jaw opening amplitude, or phase angle to a bivariate correlation analysis. None of the bivariate correlational analyses yielded a signification correlation [duration x age: *r*(34) = .163, *p* = .357; jaw amplitude x age: *r*(34) = .055, *p* = .756; phase angle x age: *r*(34) = .093, *p* = .601].

To determine if phase angles varied systematically with tongue body cycle duration or jaw amplitude, bivariate correlation analyses were conducted across all talkers as well as talkers within each group. For the correlation analyses, the normality of the parameters was assessed for the entire sample as well as for each group using the Shapiro-Wilk test. The Shapiro-Wilk test yielded significant findings for durations, jaw opening amplitudes, and phase angles when all talkers were considered (*p* ≤ .046) as well as for jaw opening amplitudes of talkers with MS, HD, and typical talkers (*p* ≤ .001), and for phase angles of CON (*p* = .025). Therefore, a Spearman correlation was conducted when considering all talkers, when correlating jaw opening amplitudes with phase angles in talkers with PD, MS, HD, and CON, and when correlating duration with phase angles in CON. Otherwise, Pearson’s correlations were conducted.

## Results

### Aim 1: Between-Group Differences in Tongue Body Cycle Durations

Estimated marginal means and standard errors of cycle durations are provided in Table 2. Fig 4 shows the boxplots for each group’s cycle durations. A significant main effect of group on cycle duration was found [*F*(4,105.265) = 19.795, *p* < .001]. A summary of the post-hoc tests can be found in Table 3. Post-hoc analyses revealed that the average cycle duration was significantly shorter in the CON group compared to the MS group (*p* < .001, mean difference = −0.082s) and ALS group (*p* < .001, mean difference = −0.131s). In addition, the PD group had also significantly shorter cycle duratio ns than the MS group (*p* < .001, mean difference = −0.107s) and the ALS group (*p* < .001, mean difference = −0.156s). Furthermore, the HD group had also significantly shorter cycle durations than the MS group (*p* = .007, mean difference = −0.076s) and the ALS group (*p* < .001, mean difference = −0.126s). No significant differences were observed between the CON group and either the PD or HD groups, nor between the MS group and the ALS group or the HD group and the PD group.

**Fig 4.**
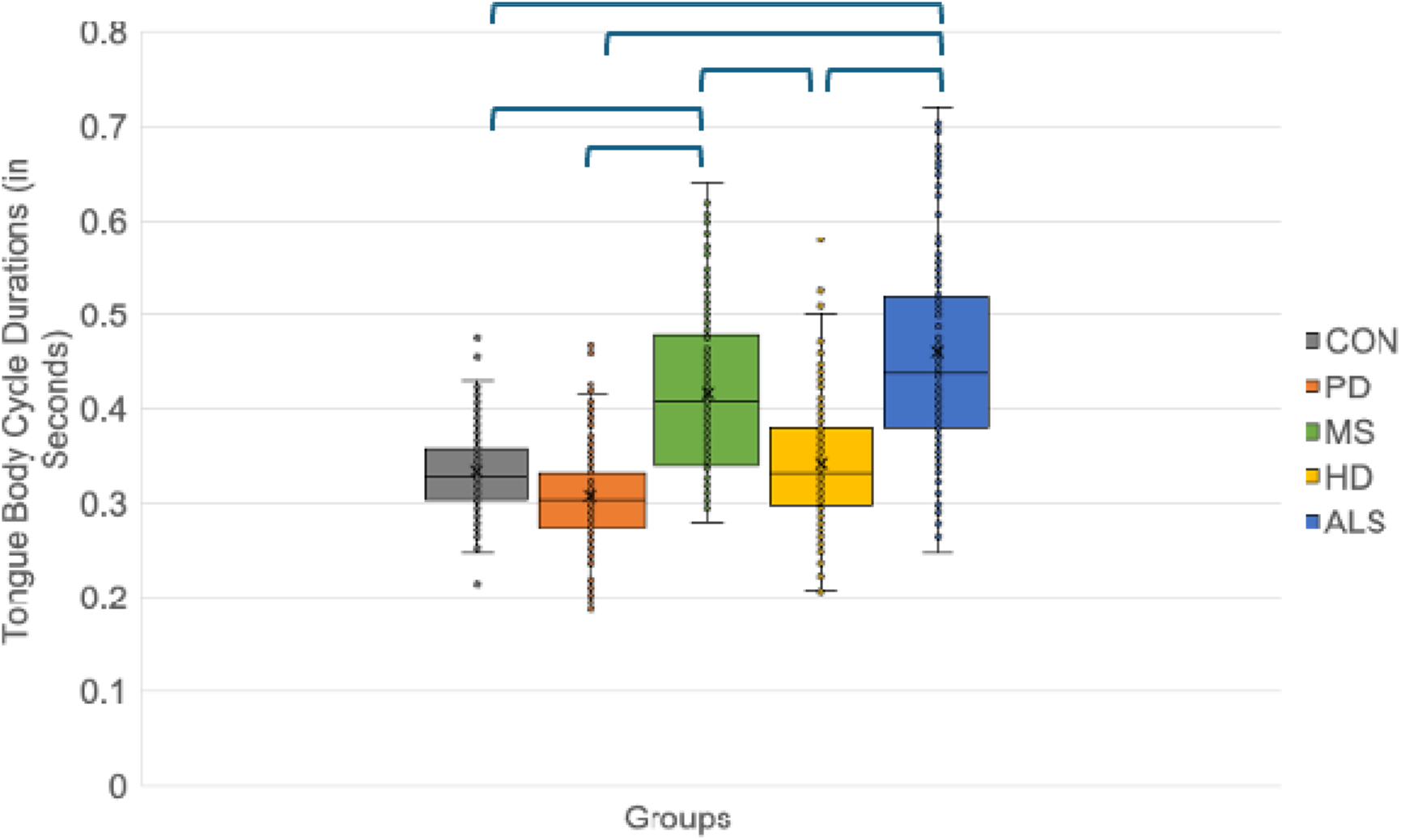
Cycle durations across groups. Brackets show significant between-group findings (*p* < .05).

**Table 2.** Estimated marginal means (*SE*) for tongue body cycle durations.

|  | CON | PD | HD | MS | ALS |
|---|---|---|---|---|---|
| Cycle duration in seconds (*SE*) | 0.334<br>(0.011) | 0.310<br>(0.013) | 0.340<br>(0.016) | 0.416<br>(0.015) | 0.465<br>(0.016) |

**Table 3.** Findings of post-hoc tests for pairwise group comparisons of tongue.

| Group Comparison | Mean Difference | *p*-Value |
|---|---|---|
| ALS vs. CON | 0.131 | <.001 |
| ALS vs HD | 0.126 | <.001 |
| ALS vs MS | 0.050 | .250 |
| ALS vs PD | 0.156 | <.001 |
| HD vs CON | 0.005 | 1.000 |
| HD vs MS | -0.076 | .007 |
| HD vs PD | 0.030 | 1.000 |
| MS vs CON | 0.082 | <.001 |
| MS vs PD | 0.107 | <.001 |
| PD vs CON | -0.025 | 1.000 |

Note. Adjustments for multiple comparisons: Bonferroni

### Aim 2: Between-Group Differences in Jaw Opening Amplitudes

Estimated marginal means and standard errors of jaw opening amplitudes are provided in Table 4. Fig 5 shows the boxplots for each group’s jaw amplitudes. A significant main effect of group on jaw amplitudes was found [*F*(4,1061) = 6.113, *p* < .001]. A summary of the post-hoc tests can be found in Table 5. Post-hoc analyses revealed that talkers with ALS have significantly larger jaw opening amplitudes than talkers with PD (*p* = .001, mean difference = 3.97mm) and typical talkers (*p* = .001, mean difference = 3.93mm). All other pairwise group comparisons were not significant.

**Fig 5.**
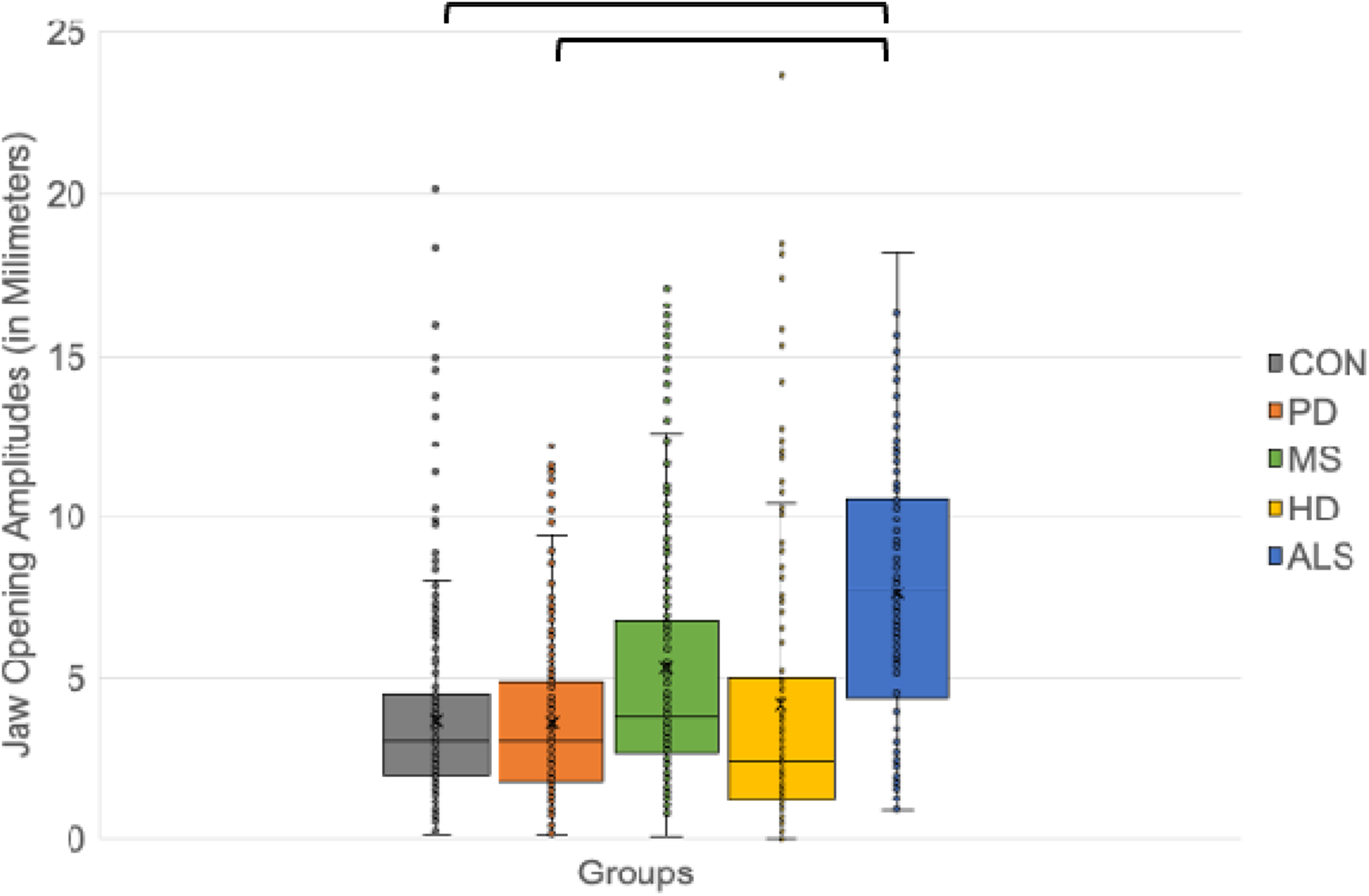
Jaw opening amplitudes across groups. Brackets show significant between-group findings (*p* < .05)

**Table 4.**
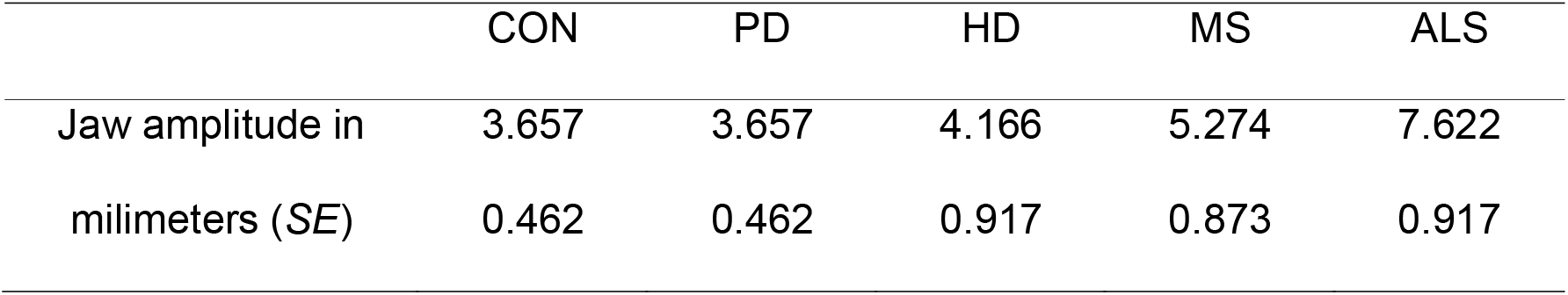
Estimated marginal means (*SE*) for jaw opening amplitudes in [kai].

|  | CON | PD | HD | MS | ALS |
|---|---|---|---|---|---|
| Jaw amplitude in milimeters (*SE*) | 3.657<br>0.462 | 3.657<br>0.462 | 4.166<br>0.917 | 5.274<br>0.873 | 7.622<br>0.917 |

**Table 5.** Findings of post-hoc tests for pairwise group comparisons of jaw amplitudes during the jaw opening movement in [kai].

| Group comparison | Mean Difference | *p*-value |
|---|---|---|
| ALS vs. CON | 3.930 | .001 |
| ALS vs HD | 3.456 | .078 |
| ALS vs MS | 2.348 | .448 |
| ALS vs PD | 3.966 | .001 |
| HD vs CON | 0.474 | 1.000 |
| HD vs MS | -1.108 | 1.000 |
| HD vs PD | 0.509 | 1.000 |
| MS vs CON | 1.582 | .611 |
| MS vs PD | 1.617 | .611 |
| PD vs CON | -0.035 | 1.000 |

Note. Adjustments for multiple comparisons: Bonferroni

### Aim 3 Testing: Between-Group Differences in Phase Angles

Estimated marginal means and standard errors of phase angles are provided in Table 6. Fig 6 shows the boxplots of each group’s phase angles. A significant main effect of group on phase angle was found [F(4,104.749) = 3.300, p = .014]. Table 7 provides a summary of all post-hoc tests. Post-hoc analyses revealed that the phase angles of the ALS group were significantly larger than the phase angles of the CON group (p = .042, mean difference = 13.562 degrees) the PD group (p = .026, mean difference = 15.294 degrees), the HD group (p = .049, mean difference = 15.483), and the MS group (p = .036, mean difference = 15.742 degrees). All other pairwise group comparisons were not statistically significant.

**Fig 6.**
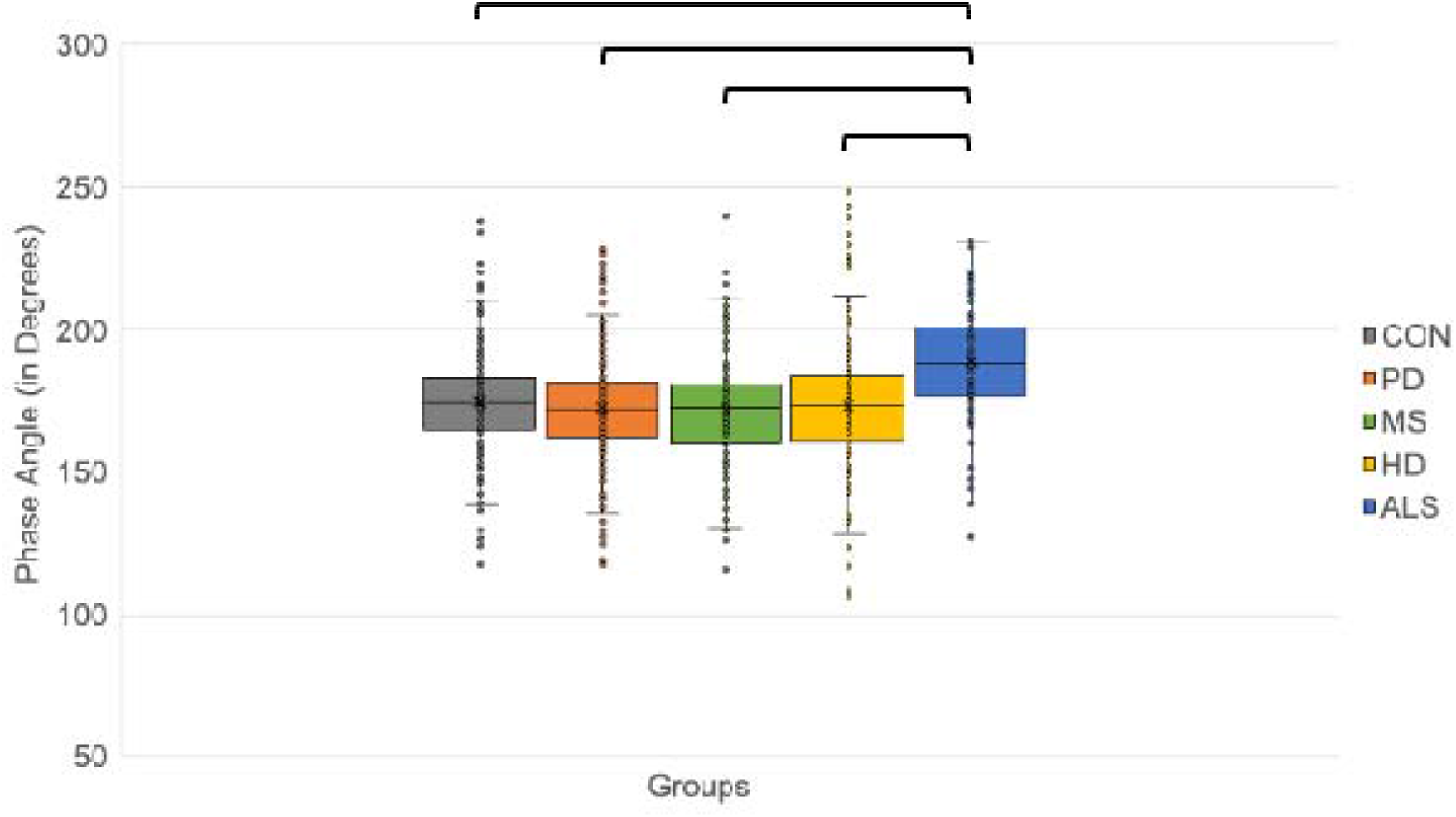
Phase angles across groups. Brackets show significant between-group findings (*p < .05)*

**Table 6.** Estimated marginal means (*SE*) for phase angles.

|  | CON | PD | HD | MS | ALS |
|---|---|---|---|---|---|
| Phase angle φ in degrees (*SE*) | 174.5<br>(2.7) | 172.76<br>(3.2) | 172.6<br>(3.8) | 172.3<br>(3.7) | 188.1<br>(3.8) |

**Table 7.** Findings of post-hoc tests for pairwise group comparisons for phase angles.

| Group Comparison | Mean Difference | *p*-value |
|---|---|---|
| ALS vs CON | 13.562 | .042 |
| ALS vs HD | 15.483 | .049 |
| ALS vs MS | 15.742 | .036 |
| ALS vs PD | 15.294 | .026 |
| HD vs CON | -1.921 | 1.000 |
| HD vs MS | 0.259 | 1.000 |
| HD vs PD | -0.189 | 1.000 |
| MS vs CON | -2.180 | 1.000 |
| MS vs PD | -0.448 | 1.000 |
| PD vs CON | -1.732 | 1.000 |

### Aim 4: Relationship between Tongue Body Cycle Duration and Jaw Phase Angle

Each talker’s phase angles and cycle durations were averaged across the ten repetitions and the resulting mean phase angles and cycle durations were submitted to a bivariate correlational analysis. Panel A of Fig 7 shows the association between the mean phase angle and the mean duration for all talkers. Table 8 displays the findings for the bivariate correlation analyses. No significant correlation was found between cycle duration and phase angles across talkers of all groups [*r*_S_(110) = .141, *p* = .141]. The coefficient of determination (R²) of 0.02 indicates that tongue body cycle duration accounts for almost none of the variation in inter-articulatory phase angles. Similarly, no significant correlation was observed within the CON group [*r*_S_(34) = .135, *p* = .445], MS group [*r*_P_(18) = .087, *p* = .733], HD group [*r*_P_(17) =.061, *p*= .815], ALS group [*r*_P_ (17) = -.134, *p* = .608], and the PD group [*r*_P_(24) = -.194, *p* = .363].

**Fig 7.**
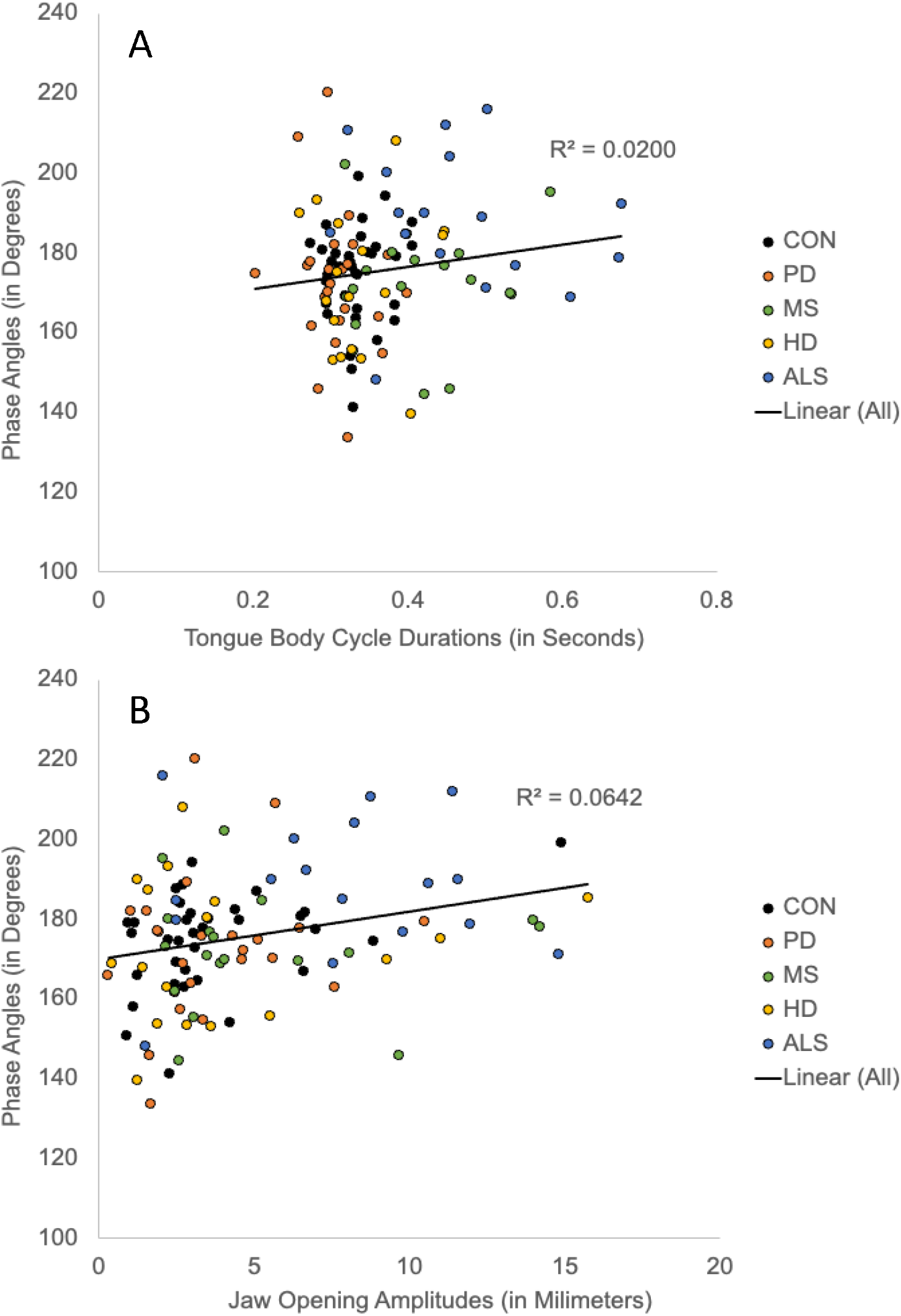
Phase angles as a function of tongue body cycle durations (Panel A) and jaw opening amplitudes (Panel B). Groups are denoted in different colors. The solid line indicates the linear regression fit across all talkers regardless of group membership.

**Table 8.**
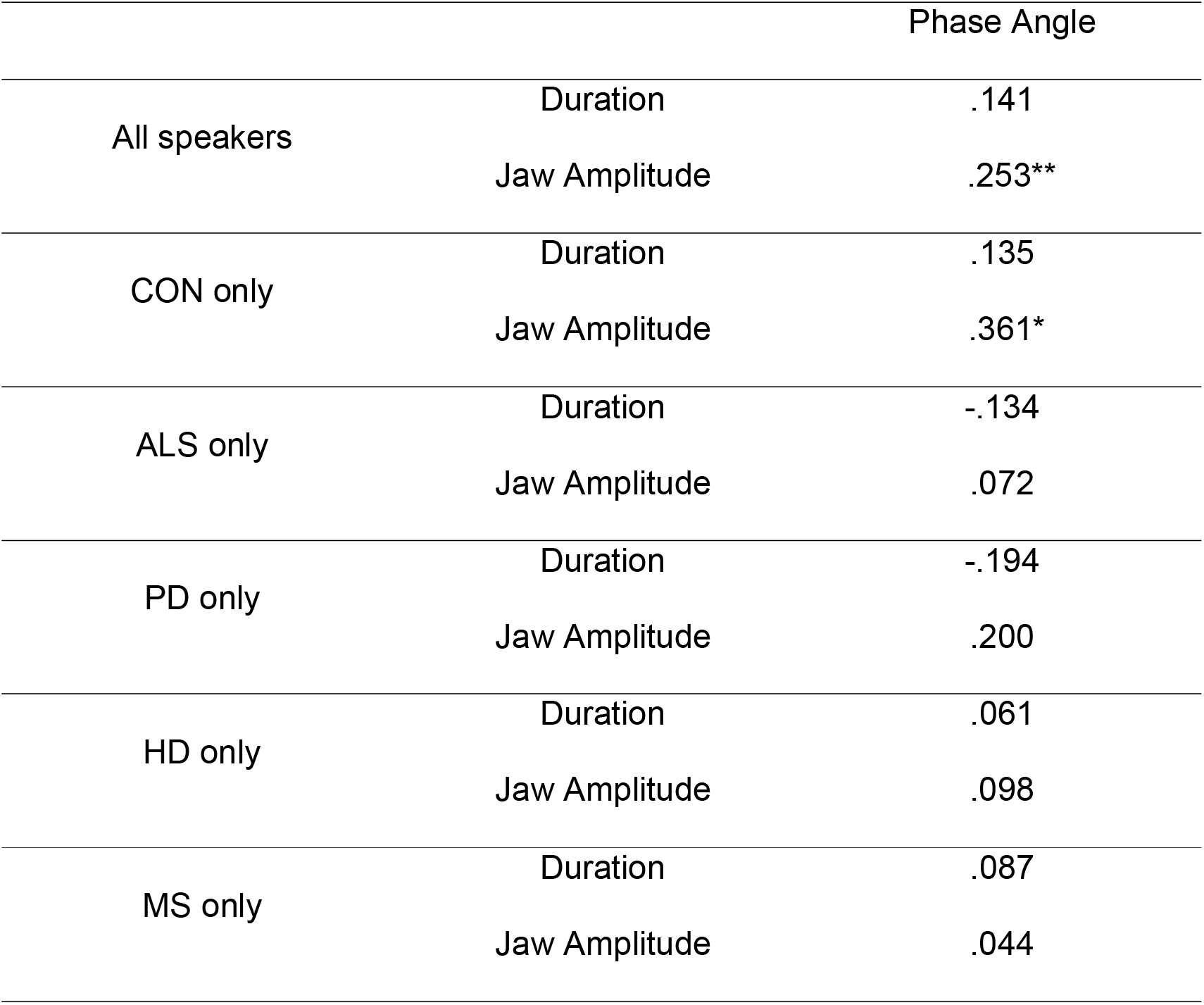
Bivariate correlations between kinematic variables and phase angle across groups and for all participants. Values represent correlation coefficients. Asterisks indicate statistical significance (* p < .05, ** p < .01).

|  |  | Phase Angle |
|---|---|---|
| All speakers | Duration | .141 |
|  | Jaw Amplitude | .253** |
| CON only | Duration | .135 |
|  | Jaw Amplitude | .361* |
| ALS only | Duration | -.134 |
|  | Jaw Amplitude | .072 |
| PD only | Duration | -.194 |
|  | Jaw Amplitude | .200 |
| HD only | Duration | .061 |
|  | Jaw Amplitude | .098 |
| MS only | Duration | .087 |
|  | Jaw Amplitude | .044 |

### Aim 5: Relationship between Jaw Opening Amplitude and Phase Angles

Each talker’s phase angles and jaw opening amplitudes were averaged across the ten repetitions and the resulting mean phase angles and cycle durations were submitted to a bivariate correlational analysis. Panel B in Fig 7 shows the association between the mean phase angle and the mean jaw opening amplitudes for all talkers. Table 8 displays the findings for the bivariate correlation analyses. A significant positive correlation was found between phase angles and jaw opening amplitudes across talkers of all groups [*r*_S_(110) = .253, *p* = .008]. However, the coefficient of determination (*R*²) of 0.064 indicates that jaw opening amplitudes accounts for only 6.4% of the variation in inter-articulatory phase angles. Furthermore, the correlational analysis only yielded a significant finding in typical talkers [r_S_(34) = .361, p = .036] indicating that the positive correlation across all talkers was mostly driven by the association between phase angles and jaw opening amplitudes in typical talkers. No significant correlations were observed within the MS group [*r*_S_(18) = .044, *p* = .861], HD group [*r*_S_(17) =.098, *p* = .708], ALS group [*r*_P_(17) = .072, *p* = .782], and the PD group [*r*_P_(24) = .200, *p* = .349].

## Discussion

This study investigated inter-articulatory timing patterns of the tongue body and jaw in talkers with different neurological conditions (i.e., PD, HD, MS, and ALS). First, we tested for between-group differences in tongue body cycle durations and jaw opening amplitudes. Findings for between-group differences in tongue body cycle duration aligned with previous reports of acoustic studies on speaking rate abnormalities in talkers with PD, ALS, MS and CON [e.g., 39–41]. That is, talkers with ALS and MS had significantly longer tongue body cycle durations indicating that their speaking rates were slower than those of talkers with PD and CON. Findings of comparable tongue body cycle durations in talkers with HD and CON differed from previous reports of prolonged segment durations and slowed speaking rates in talkers with HD [37,38]; however, they aligned with reports of a dysarthria subgroups of talkers with HD with an unaffected speaking rate [36]. Furthermore, findings for jaw opening amplitudes were congruent with previous kinematic studies [42]. That is, talkers with ALS had significantly larger jaw opening amplitudes than talkers with PD and CON and tended to be larger than those of talkers with MS and HD.

With regards to inter-articulatory timing patterns, we examined phase angles and expected them to be smaller or larger than 180 degrees in talkers with HD, MS, and PD due to less synchronized movements given that their neuropathologies involve impairment of the basal ganglia or cerebellar control circuits. In addition, phase angles of talkers with ALS were expected to be close to 180 degrees given their potential overreliance on the jaw and limited independent tongue movements during speech. However, our findings did not support this hypothesis. We found that phase angles of talkers with ALS were significantly greater than those of talkers with MS, PD, HD, and typical talkers. Although talkers with ALS produced abnormally large jaw movements, they did not exhibit more synchronized tongue and jaw movements than talkers with MS, PD, HD, or typical talkers. Specifically, their phase angles were on average 8.4 degrees above 180 degrees while talkers with HD, MS, PD and typical talkers were on average between 5.3 degrees (talkers with HD) and 8.3 degrees (talkers with MS) below 180 degrees. A weak positive correlation was observed between jaw opening amplitudes and phase angles when considering all talkers and when considering only typical talkers. This finding suggests that the size of the jaw movement cannot be the main driver of the between-group differences in phase angles, especially because the weak positive association between jaw opening amplitudes and phase angles observed across all talkers was predominantly driven by the articulatory performance of typical talkers, not by talkers with ALS.

Alternatively, we examined if phase angles vary with speech rate abnormalities across these clinical groups given previous findings of speech rate effects on phase angles in typical talkers. That is, talkers with MS and ALS, and potentially also talkers with HD, were expected to differ in their phase angles from talkers with PD and typical talkers due to their speech rate differences. However, our findings did not support this hypothesis. Only talkers with ALS but not talkers with MS differed in their phase angles from those of talkers with PD and typical talkers. No significant association between cycle duration and phase angles were observed when considering all talkers or when considering only talkers within the same group. Our findings highlight an important distinction between habitual speaking rate differences and experimentally elicited rate manipulations. While previous studies demonstrated that cued changes in speaking rate (e.g., instructing a speaker to speak faster or slower) can systematically alter phase angles, our results indicate that habitual cycle durations do not exert the same driving effect. Specifically, despite significant differences in habitual tongue body cycle durations across clinical and control groups, no significant associations between cycle duration and phase angles were observed. This suggests that a speaker’s intrinsic, habitual rate, whether typical or slowed due to pathology, does not inherently constrain inter-articulatory timing in the same manner as a targeted rate adjustment paradigm.

### Disease-specific inter-articulatory timing pattern in talkers with ALS

Talkers with ALS had significantly greater phase angles than talkers with HD, MS, PD, and typical talkers. On average, phase angles of talkers with ALS were greater than 180 degrees, which indicates that the tongue body commonly reached the positional minimum at the diphthong onset before the jaw. In fact, only five of the 17 talkers with ALS (29%) did not follow this pattern and had phase angles smaller than 180 degrees. By contrast, the average phase angles of typical talkers, talkers with MS, PD, and HD were smaller than 180 degrees, which indicates that the jaw commonly reached the target before the tongue body. More specifically, only ten of the 34 CON (29%), two of the 18 talkers with MS (11%), five of the 24 talkers with PD (21%), and six of the 17 talkers with HD (35%) did not follow this pattern and had phase angles that were greater than 180 degrees. Thus, with regards to the order in which the articulators reached the target, the inter-articulatory timing patterns were fairly robust within each group.

It is important to point out that because the current study used the tongue body as the reference articulator and not the jaw, phase angles should be smaller at longer tongue body cycle durations to align with previous findings on cued slow speech effects [13]. That is because phase angles below 180 degrees indicate that the jaw leads the tongue when the tongue body is the reference (current study) whereas phase angles above 180 degree indicate that the jaw leads the tongue when the jaw is the reference sensor (previous studies). Therefore, the observed phase angles of most talkers with ALS in the current study do not align with the inter-articulatory timing pattern previously observed for cued slow speech in typical talkers.

The discrepancies between the findings for phase angles under cued slow speech in typical talkers and those for phase angles of talkers with ALS who spoke at their habitual albeit abnormally slow rate suggest that the absolute movement duration, or the absolute speech rate, may not determine a talker’s inter-articulatory timing pattern. This notion is supposed by our correlational analysis of tongue body cycle durations and phase angles, which did not yield a significant finding. Thus, it may rather be the speech task (i.e., the execution of a cued slow speech rate) that determines the inter-articulatory timing pattern. Indeed, previous studies revealed speech rate effects on phase angles in typical talkers [8,13,15]. However, in these studies, talkers were cued to increase or reduce their speaking rate. In the current study, talkers were asked to speak at their habitual rate, which resulted a wide range of tongue body cycle durations across talkers due to their dysarthria. Especially talkers with MS and ALS had significantly longer tongue body cycle durations than CON, talkers with PD, and talkers with HD. However, the absolute tongue body cycle duration, which indirectly characterized a talker’s speech rate, was not predictive of their inter-articulatory timing pattern. To better understand the unique inter-articulatory timing pattern of talkers with ALS, future studies could examine if speaking rate modifications elicit changes in the phase angles of talkers with ALS. Such insights would help determine if talkers with ALS are constrained to their specific inter-articulatory timing pattern due to their disease or if they are able to modify their inter-articulatory timing patterns in response to various speech task demands.

### Similar inter-articulatory timing patterns across talkers with MS, PD, and typical talkers

No significant differences in phase angles were found between typical talkers, talkers with MS, HD, and PD. The lack of significant differences in inter-articulatory timing patterns between typical talkers and talkers with PD and MS contradicts findings of disrupted timing patterns of limb motor control studies [43–45], whereas the lack of group-level differences in HD may be attributable to high individual variability which will be discussed later on. The findings of the current study may suggest that the speech motor system is spared from the negative impact of the basal ganglia and cerebellar neuropathologies while the limb motor system is affected. However, it is also possible that most of the talkers with dysarthria in the current study were too mild in their dysarthria severity to observe disrupted timing patterns within the speech motor system. For example, Bartle and colleagues [23] reported that only the most severe talkers with dysarthria showed disrupted tongue-jaw timing patterns.

Therefore, we compared the data of the talkers with moderate and severe dysarthria to those with mild dysarthria within each group; however, no systematic pattern could be detected. The four panels of Fig 8 illustrate the lack of a relationship between SIT scores and phase angles. However, given the limited number of talkers with moderate and severe dysarthria in each clinical group, future studies are needed to investigate timing patterns of limb and speech movements side by side and include more individuals with moderate and severe dysarthria to better understand the impact of basal ganglia and cerebellar pathologies on timing patterns across the speech and limb motor system.

**Fig 8.**
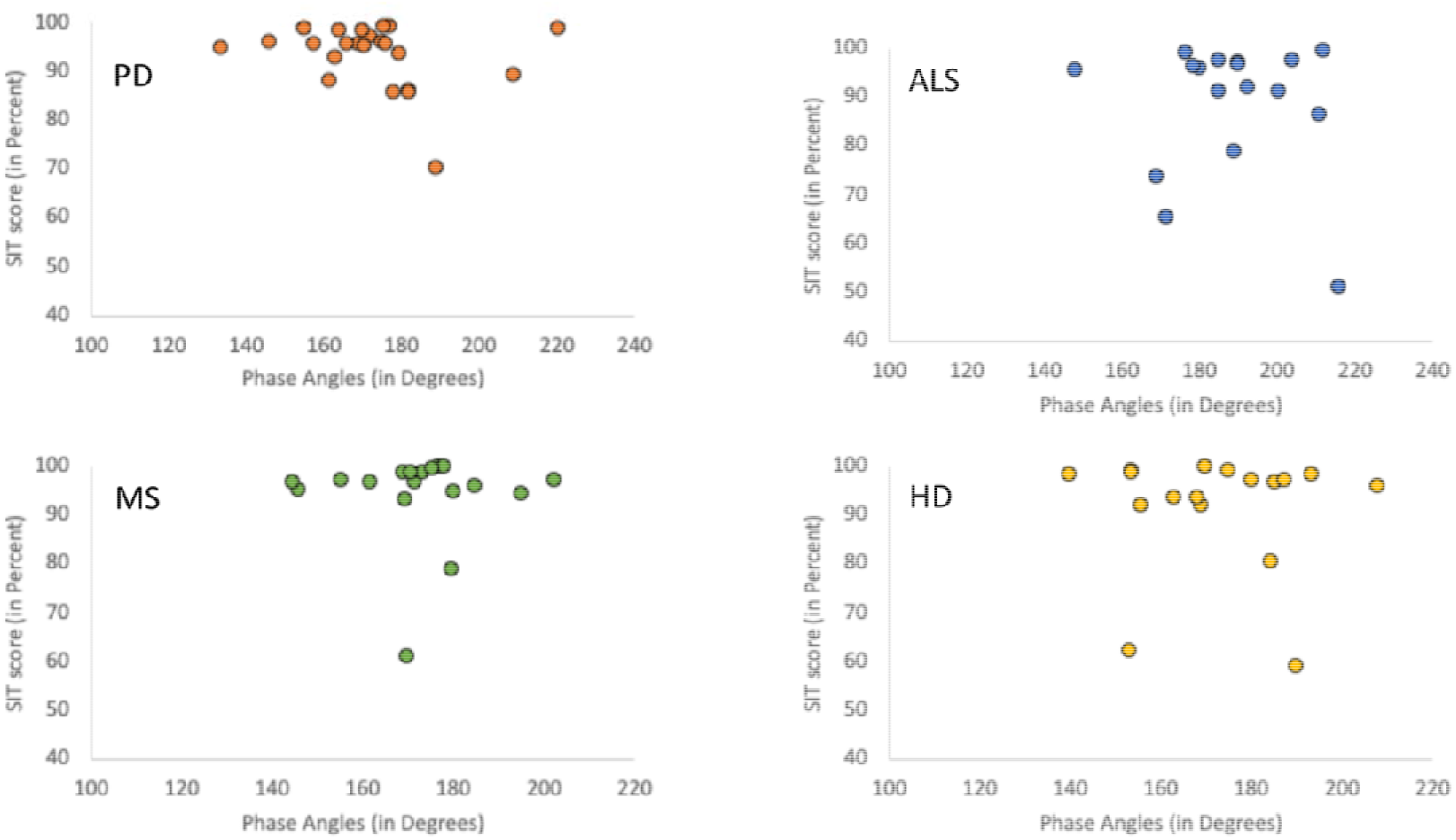
Associations between SIT scores and phase angles for each clinical group.

### Highly variable inter-articulatory timing patterns in talkers with HD

Although not evident in the group level plots, we would like to point out differences in inter-articulatory timing patterns at the individual performance level. Specifically, we noted that phase angles varied greatly across repetitions within each talker with HD.

For example, the phase angle’s coefficient of variation across trials was on average 11.4 degrees for talkers with HD whereas the coefficient of variation for talkers with PD, MS, ALS and typical talkers was on average much lower (3.9 degrees in ALS, 6.4 degrees in MS, 5.8 degrees in PD, and 7.5 degrees in CON). This observation suggests that although phase angles of talkers with HD did not significantly differ from the other clinical groups and typical talkers, the basal ganglia pathology may negatively impact the ability to produce a consistent inter-articulatory timing pattern in talkers with HD.

### Quantifying inter-articulatory timing patterns with phase angles

In the current study, we chose to characterize inter-articulatory timing patterns by measuring phase angles. Phase angles are expressed relative to 360 degrees regardless of the duration of the cycle and are heavily influenced by the shape of the movement trajectory towards and away from the target position (i.e., maximum opening position). However, it should be noted that phase angles do not provide the same insights as the latency measure (i.e., temporal lag). Although a greater latency is typically associated with a phase angle that is much smaller or larger than 180 degrees, a specific phase angle can have slightly different latencies and a specific latency can have slightly different phase angles. That is because the shape of the movement trajectory of the articulator that serves as the reference signal impacts the phase angle more than the latency itself. For example, if the shape of the tongue movement trajectory is characterized by a plateau around the maximum opening position at the onset of the diphthong [ai], phase angles will be closer to 180 degrees even if there is a relatively small lag between the jaw and tongue. By contrast, if the movement trajectory of the tongue is characterized by more “peakedness” towards and away from the maximum opening position, relatively small lags between the tongue and jaw can produce phase angles that are disproportionally smaller or larger than 180 degrees.

Due to these concerns, we informally tested the strength of association between latencies and phase angles by extracting the lag durations between the tongue and the jaw reaching the maximum opening position at the onset of the diphthong [ai] together with the phase angles across all repetitions of all talkers. Each talker’s mean latencies and phase angles were submitted to Pearson’s correlational analysis. We found a strong positive correlation of r(110) = .88, p < .001 between latencies and phase angles across talkers and similarly strong correlations within each group [PD: r(24) = .94, p < .001, HD: r(17) = .85, p < .001; MS: r(18) = .88, p < .001; ALS: r(17) = .86, p < .001; CON: r(34) = .92, p < .001]. Thus, the reported phase angles in the current study can provide good insights in the extent to which tongue and jaw movements are synchronized.

### Study limitations and future directions

Previous studies investigated inter-articulatory timing patterns by expressing the timing of the articulator of interest (tongue, upper lip, lower lip) with respect to the jaw cycle . In the current study, the tongue cycle was taken as reference to which the timing of the jaw reaching the maximal opening of the jaw at the onset of the diphthong /ai/ was expressed, which limits the comparability to previous studies. However, with cycle durations of typical talkers in the current study being comparable to those of a slow speech condition in previous studies, the phase angles of our typical talkers were well-aligned with those of previous studies when accounting for the different reference articulator [8,13]. That is, the average phase angle for our control group was about 172 degrees indicating that the jaw led the tongue body. The average phase angle of ten talkers in Masapolo and Nittrouer’s study [8] was about 200 degrees, also indicating that the jaw led the tongue tip in typical talkers.

Mathematically, phase relations measure relative spatiotemporal coupling regardless of which articulator is assigned as the reference frame. Therefore, we believe that our approach of using the tongue body as the reference was well-suited to study inter-articulatory timing patterns, specifically to explore the jaw-tongue coupling patterns. However, future studies should investigate inter-articulatory timing patterns with the traditional approach of using the jaw as the reference signal to replicate and further extend current study findings on talkers with dysarthria.

Furthermore, a major difference in the experimental design between the current and previous studies was that the target utterance did not contain two stop consonants, such as [tad, bab, bak, tak], but [kai]. However, the offset of the diphthong [ai] at the end of the tongue body cycle also required a vocal tract constriction that was consistently executed by the tongue body. Although, the jaw was consistently lowered to produce the maximum opening position at the onset of the diphthong [ai], it was not consistently raised during the diphthong. However, the jaw was always elevated after reaching the maximum opening position at the onset of the diphthong [ai] and eventually supported the constriction of [k] in “kite”. Nevertheless, due to the presence of reliable landmarks for the tongue body and jaw to reach the maximum opening position and initiate the upward movement towards vocal tract constriction at the offset of the diphthong [ai], we argue that the sequence [kai] was also well-suited to study inter-articulatory timing patterns.

Another major difference between the current study and previous studies is that we could not decouple the jaw from the tongue. Decoupling jaw movements from the posterior tongue is challenging because the jaw movements are recorded more anteriorly in the vocal tract where, due to jaw rotation, the movement amplitudes of the jaw are larger than at the location of the posterior tongue. A linear subtraction of jaw movement from the posterior tongue, which is an approach commonly used to decouple the lower lip or the tongue tip from the jaw, would not yield accurate data of independent posterior tongue movement [36]. Nevertheless, the timing of the tongue and jaw reaching the maximum opening position at the onset of the diphthong [ai] was asynchronous in most cases (983 out of 1066 productions, 92%) indicating that independent tongue movements are reflected in the tongue movement trajectory. However, the shape of the tongue movement could be partially affected by the jaw in the current study and this could impact the specific value of the phase angle. However, it would not affect whether a phase angle was smaller or larger than 180 degrees because the order of the articulators reaching the target and not the specific tongue position and velocity determines whether a phase angle is smaller or larger than 180 degrees (i.e., if the jaw is reaching the target before the tongue, the phase angle is smaller than 180 degrees if the tongue body is the reference signal). Therefore, the overall finding of the current study, which suggest that talkers with ALS differ in their inter-articulatory timing patterns from talkers with PD, MS, HD, and typical talkers was primarily driven by the order of the articulators reaching the maximum opening position at the onset of the diphthong [ai]. Nevertheless, it is important to develop a decoupling algorithm that can accurately determine independent posterior tongue movements for future studies to replicate the current findings.

In the current study, we did not control for medication status in talkers with HD and surgical intervention (DBS) in talkers with PD. However, we did not observe any systematic differences in phase angles due to the medication status in talkers with HD. One talker with HD on medication produced the largest phase angles of the HD group while two other talkers with HD on medication had very small phase angles and the remaining three talkers with HD on medication fell somewhere in the middle of the group’s distribution. Similarly, two of the three talkers with PD who had undergone DBS treatment had phase angles comparable to those of talkers with PD who did not have DBS. The third talker with DBS produced the largest phase angles of all talkers with PD. Thus, no systematic pattern could be detected that suggest medication status or DBS could be considered confounding factors.

Finally, future studies are needed to examine inter-articulatory timing patterns across a wide range of phonetic contexts to determine if current findings generalize or if they are stimulus-specific. Future studies should also include the tongue tip and jaw as well as the upper lip and lower lip or jaw as articulators of interest to determine if inter-articulatory timing patterns reveal similar disease-related group differences.

## Conclusion

This study investigated temporal, spatial and spatiotemporal alterations between tongue body and jaw movements across several diseases. While tongue body cycle durations were significantly prolonged in ALS and MS, jaw amplitudes were enlarged specifically in ALS. Inter-articulatory phase angles differentiated ALS from all other cohorts. Talkers with ALS exhibited significantly larger phase angles compared to PD, HD, and MS groups, whose phase angles remained comparable to typical talkers. This highlights distinct spatiotemporal timing profiles in ALS that evolve largely independently of speech rate or jaw amplitude variations.

## Acknowledgments

We thank all participants for their willingness to take part in this study, Daniel Kim and Claudia Raines for their help with the data collection and Angela Ding for assisting with the data analysis.

## Data Availability Statement

The data used to support this study’s findings are available as supplementary material.

